# Continuous Outcome Estimation in N-of-1 Trials for Accelerated Decision-Making

**DOI:** 10.1101/2025.02.05.25321704

**Authors:** Victoria Defelippe, František Bartoš, Eric-Jan Wagenmakers, Kees P.J. Braun, Floor E. Jansen, Willem M. Otte

## Abstract

N-of-1 trials aim to determine the therapeutic effect for a single individual. This individualized approach necessitates collecting multiple data points over time through repeated alternating periods of active treatment and a comparator or control condition. The extended duration of the treatment periods may increase patient burden, prolong placebo exposure, and increase the likelihood of study discontinuation. In theory, treatment responders (or non-responders) can be identified early during the trial if the therapeutic effect is strong (or completely lacking). There are no theoretical constraints to evaluate treatment efficacy more regularly – not only after a predetermined number of treatment periods – given that the individualized character of the N-of-1 study permits a statistical model update as soon as new data becomes accessible. Regularly updating estimates on treatment effects allows clinicians to accelerate clinical decision-making regarding N-of-1 study termination. This study examines the importance of continuous treatment effect estimation in N-of-1 trials through simulation and re-analysis of existing trial datasets of neurological diseases. Results indicate that treatment efficacy decisions can be expedited when outcome estimation is performed continuously rather than delayed until the end of the trial.

## Introduction

An N-of-1 trial is characterized by a crossover design within a single subject who undergoes multiple cycles of alternating placebo or verum treatment periods (Irwig et al. 1995). The primary goal of a clinical N-of-1 approach – from the perspective of care improvement – is to establish treatment efficacy for an individual subject without attempting to extrapolate findings to the broader population with the same condition (Defelippe et al. 2023; Fountzilas et al., 2022). Establishing such a contextual and subject-specific assessment of therapeutic efficacy necessitates the collection of multiple data points. Therefore, N-of-1 trials are particularly well-suited for investigating chronic, neurological conditions characterized by measurable symptoms or recurrent paroxysmal events (Defelippe et al., 2024; Stunnenberg et al., 2022; Margolis et al., 2019). The occurrence of recurrent events provides a unique opportunity for conducting robust statistical analyses of data collected within N-of-1 trials. However, obtaining sufficient data points to assess individual-level efficacy requires extended observation periods and multiple crossovers, potentially resulting in longer trial duration compared to traditional randomized controlled clinical trials. Extended trial duration presents risks of increased patient burden and study attrition while prolonging subject exposure to either placebo (control) treatment or potentially ineffective verum interventions.

An N-of-1 trial can be completed once there is sufficient evidence to establish either the efficacy or inefficacy of the verum intervention. Evaluations of therapeutic efficacy are usually conducted either following the completion of each treatment cycle or after a predetermined sequence of verum and placebo periods. Theoretically, the requisite number of verum and placebo periods for assessing the therapeutic efficacy may extend to whatever quantity is statistically necessary to achieve robust estimation parameters. In practice, however, N-of-1 trials are generally constrained to a limited number of cycles, primarily to facilitate and maintain patient adherence to the protocol. This methodological constraint exemplifies the tension between optimizing patient compliance and maintaining statistical rigor and relevance.

Beyond the conventional approach of analyzing efficacy after discrete treatment periods or upon trial completion, the Bayesian framework allows the possibility to evaluate the treatment effect continuously, whenever new data becomes available. This methodology permits iterative updating of results – thereby generating evidence of efficacy or its absence – at customizable temporal intervals: daily, weekly, or monthly. Such continuous evaluation methodology could be operationalized clinically through digital data collection methods via a smartphone App. Consequently, the N-of-1 trial’s duration might be completed earlier because new data is analyzed as it emerges. This methodological refinement holds the potential to minimize both economic costs and patient burden.

The empirical value of continuously treatment effect updating within N-of-1 trial protocols remains to be quantified. Therefore, this study investigates the potential benefits of implementing continuous data updating protocols. Here we evaluate the significance of continuous treatment effect estimation through simulation of an N-of-1 trial representative of refractory epilepsy cases, and through re-analysis of three published neurological N-of-1 trials encompassing epilepsy and severe headache conditions, comparing outcomes with standard evaluation protocols.

We hypothesized that continuous outcome estimation, as opposed to discrete post-cycle evaluations, facilitates accelerated decision-making in N-of-1 trials.

## Methods

The present investigation comprises two methodological components: first, a simulated N-of-1 trial protocol, and second, a comprehensive re-evaluation of three previously conducted N-of-1 trials. The simulated and reconstructed data and analysis scripts (R language) are available online (GitHub).^1^

### 1. Simulated N-of-1 trial data

We simulated an N-of-1 trial in a patient with severe epilepsy characterized by frequent seizure activity (mean frequency: 5 seizures per day) (**Figure 1**). The protocol encompassed data analysis from five consecutive treatment cycles, each including one placebo and one verum period. Data is collected from daily seizure reports. A pre-intervention baseline period served as a comparative reference standard. The verum intervention was parameterized with a net seizure reduction effect of 0.5, derived from a total verum effect of 0.6 minus a placebo effect of 0.1. Seizures were simulated utilizing a Poisson distribution model (generating positive integers), selected for its established appropriateness in characterizing clinical seizure frequency patterns.

**Figure 1.**
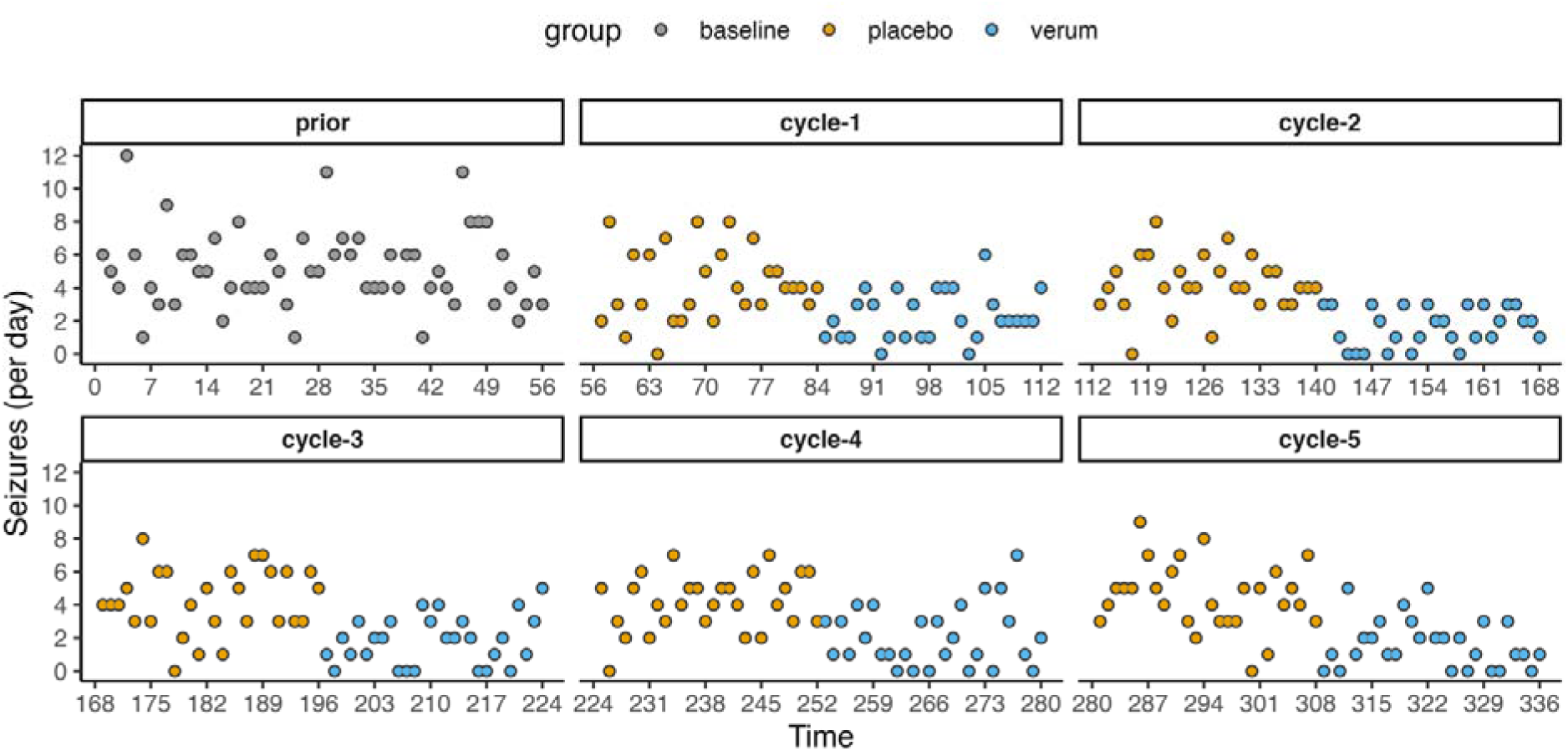
Simulated seizures in refractory epilepsy patient during baseline and five consecutive cycles with a placebo and verum period.

We modeled the daily seizure outcome with a Bayesian generalized linear model (logarithmic link function; Poisson probability distribution (baseline Poisson rate: 5; ‘brms’ R package). We updated the model after each day, starting on the day when at least one placebo and one verum datapoint is available.

Two competing models were specified: H1, which includes the period variable as a predictor, and H0, which replaces the period variable with a null predictor. The H1 model incorporated three period levels (‘baseline’, ‘placebo’, and ‘verum’), while the null model specified two levels (‘baseline’ and ‘post-baseline’).

Truncated normal priors were defined for the intercept and slope to ensure reasonable bounds within clinically plausible seizure loads. Specifically, the intercept prior followed a normal distribution with a mean of 0 and standard deviation of 100, truncated between −10 and 10. The group difference prior also followed a normal distribution with the same parameters, truncated between −5 and 5. Both models were fit with 50,000 iterations and a 3,000-iteration warmup (MCMC).

We analyzed treatment effects in two ways:

1. model comparison was conducted using Bayes factors (**Box 1**). The Bayes factors were obtained using bridge sampling (Gronau et al., 2020) and graded the evidence for including the period variable. The analysis featured two rival models: H1 (positing a difference between verum and placebo) and H0 (assuming equivalence between verum and placebo). The relative evidence strength reported using the logarithm of the Bayes factor, where higher values indicate stronger evidence that the observed data’s generative process aligns with H1 rather than H0. Following established conventions (Jeffreys, 1961; Wagenmakers et al., 2011), the continuous logarithmic Bayes factor was categorized into interpretative grades: negative values indicate evidence favoring H0 over H1, while positive values are classified as follows: “<0”: ‘evidence for H0 rather than for H1’, “<1.1”: ‘anecdotal evidence for H1’, “<2.3”: ‘substantial evidence for H1’, “<3.4”: ‘strong evidence for H1’, “<4.6”: ‘very strong evidence for H1’, and “>4.6”: ‘extreme evidence for H1’.
2. Quantification of the minimally clinically important difference (MCID) using the posterior probability of a treatment effect (**Box 2**). Whereas a commonly used MCID in epilepsy is 50% seizure reduction, in refractory epilepsy 30% has also been accepted by regulators as justifying treatment continuation and reimbursement (Staatscourant, 2022). The MCID used for clinical decision making can be patient or syndrome specific. In this simulation we used a MCID of 30%. We quantified and plotted the 30% seizure reduction certainty over time at each update of the posterior probability distributions. We compared these patterns with the certainties calculated at the standard evaluation time points (end of each cycle), to determine if less time is required to reach a similar level of certainty of estimated and sustained efficacy, given the Bayes Factor shows relative evidence in favor of H1 (verum effect) (**Figure 2**).

**Figure 2.**
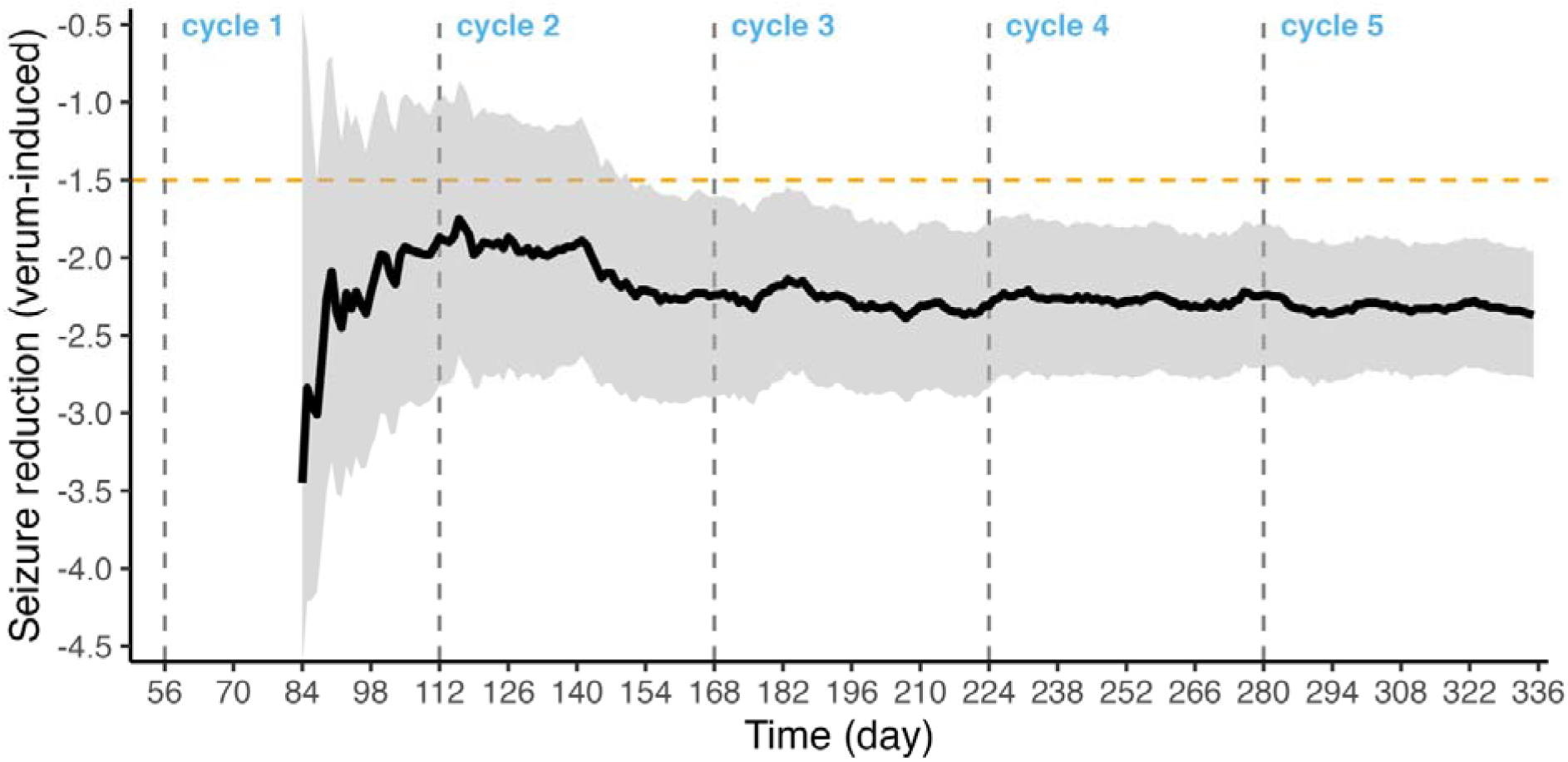
95% credibility interval for the verum-induced seizure reduction (placebo-corrected) for each day with simulated refractory seizures (Figure 4). The minimal clinically meaningful reduction of 30% is depicted in orange with a horizontal line (i.e., 1.5 seizures less). During 3/5 of the second cycle – at day 152 – the credibility interval falls below the minimally clinically relevant threshold of 30% (i.e., median daily seizure reduction: 2.21, 95% credibility interval: 1.51 to 2.92).

#### Box 1.

**Bayes factor.**

In the methodological framework of clinical trials, the Bayes factor (BF) emerges as a sophisticated statistical metrics for comparing competing hypotheses, offering a nuanced alternative to traditional frequentist approaches. The BF, expressed as BF□□ = p(D|H□)/p(D|H□), where p(D|H□) represents the likelihood of the observed data under the alternative hypothesis and p(D|H□) under the null hypothesis, quantifies the relative evidence supporting one hypothesis over another.

In the context of group comparisons, this becomes particularly salient when evaluating treatment efficacy, where BF > 1 indicates evidence favoring the alternative hypothesis (i.e., the presence of a treatment effect), while BF < 1 supports the null hypothesis. The interpretation follows a calibrated scale where, for instance, 3 < BF□□ < 10 suggests substantial evidence, and BF□□ > 10 indicates strong evidence for the alternative hypothesis.

In this study we used logarithmic Bayes factors because its transforms the multiplicative nature of the BF into an additive scale, simplifying the assessment of evidence strength. Specifically, a log BF > 0 corresponds to evidence favoring the alternative hypothesis, with higher positive values indicating stronger support. Conversely, a log BF < 0 suggests evidence supporting the null hypothesis. This logarithmic approach also mitigates issues with extreme Bayes factors, making the results more robust and easier to report.

Crucially, and unlike p-values, the Bayes Factor allows for the accumulation of evidence in favor of the null hypothesis and provides a more intuitive framework for interim analyses in adaptive trial designs, though it necessitates careful specification of prior distributions that reflect pre-existing knowledge or theoretical constraints.

#### Box 2.

**Posterior Probability and MCID.**

In the analytical framework of clinical research, posterior distributions offer a comprehensive probabilistic representation of parameter estimates following the integration of prior knowledge with observed data, fundamentally adhering to the equation P(θ|D) ∝ P(D|θ)P(θ), where P(θ|D) denotes the posterior distribution, P(D|θ) the likelihood, and P(θ) the prior distribution.

The Minimal Clinically Important Difference (MCID), when contextualized within this Bayesian paradigm, can be evaluated through the posterior probability that the treatment effect exceeds a predetermined threshold δ, expressed as P(θ > δ|D). This probabilistic interpretation becomes particularly illuminating when examining the entire distribution of plausible treatment effects, rather than merely point estimates. The posterior distribution enables researchers to make direct probability statements about parameters of interest, such as “the probability that the treatment effect exceeds the MCID is 0.95,” providing a more nuanced and clinically interpretable framework for decision-making.

This approach proves especially valuable when considering heterogeneity in treatment responses and when determining whether observed differences not only achieve statistical significance but also reach the threshold of clinical meaningfulness, thereby bridging the often-encountered gap between statistical and clinical significance.

### 2a. Re-evaluating a previous N-of-1 trial: nicotine treatment for epilepsy

A classic example of a successfully conducted N-of-1 trial in a person with refractory epilepsy is presented by Willoughby et al., 2003. Nicotine patches reduced the seizure load (one or more per day) to zero (mean: 0.01). The N-of-1 trial was preceded by an open-label period with daily seizure reporting. A follow-up double-blinded N-of-1 trial, with placebo and nicotine patches – each used for three periods of 2 weeks – provided unbiased evidence. The original seizure diary and seizure data for both open-label and N-of-1 trial periods (provided in the original publication) were reconstructed into a sequential plot (Supplementary material **Figures S1** and **S2**). **Figure 3** provides the N-of-1 trial data only.

**Figure 3.**
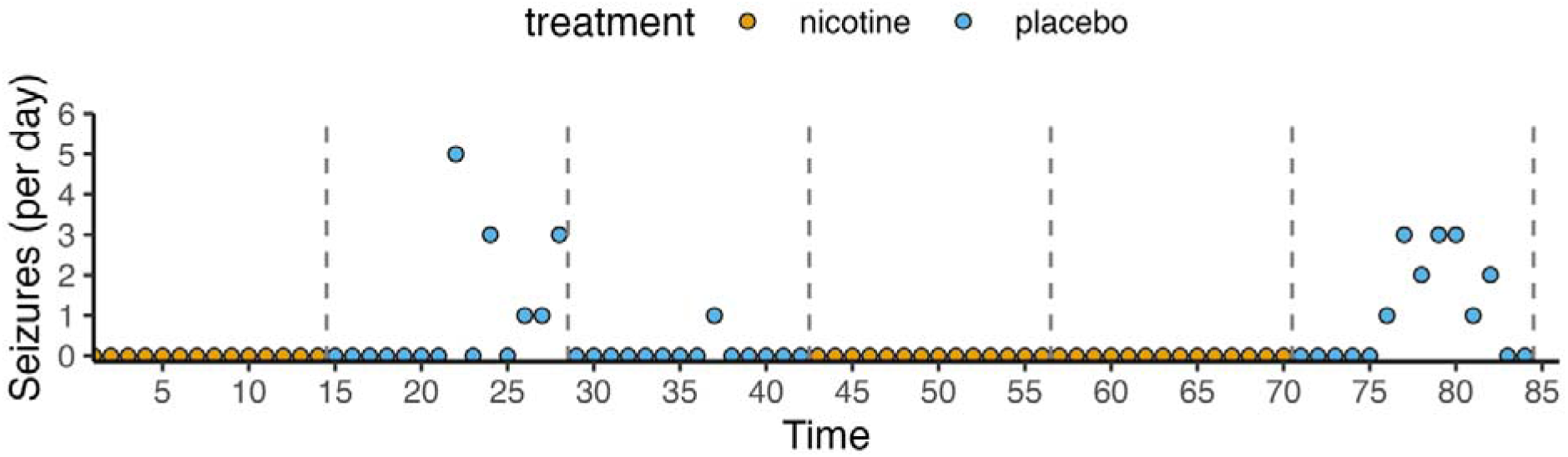
The N-of-1 trial daily seizure recording data was reconstructed from Willoughby et al., 2003 with three cycles consisting of two periods of two weeks using nicotine (n) or placebo patches (p) (12 weeks; 84 days) (design: n-p-p-n-n-p). Not a single seizure was recorded during nicotine treatment. The highest daily seizure count in the placebo period was five.

Unlike in the previous simulation, no explicit minimum clinically important difference (MCID; e.g., a seizure reduction of at least 30% or more) was reported in the original N-of-1 trial. Therefore, we focused on the effect existence rather than the smallest improvement considered worthwhile by a patient. An informative index for the effect, assuming the effect is present, is the direction probability (Makowski et al., 2019). The probability of direction ranges from 50% to 100%, representing the certainty with which an effect is positive or negative (i.e., seizure frequency reduction or increase). The direction probability is solely based on posterior distribution and does not assess the magnitude nor the significance of the effect. It is also strongly correlated with the frequentist (one-sided) p-value (Wagenmakers et al., 2023). Similarly, the p-value and probability of direction do not provide information in favor of the probability of no effect.

We also characterized the logarithmic Bayes factor after the first and second periods. We modeled the seizure outcome with a Bayesian generalized linear model and priors (i.e., a truncated normal with a range of −10 to 10 for the intercept and −5 to 5 for the period differences) comparable to the model used for the simulated seizures.

### 2b. Re-evaluating a previous N-of-1 trial treatment for prolactinoma-related headache

A second continuous N-of-1 trial reevaluation examines data from a 38-year-old female participant in a four-day study. Williams and colleagues (1986) assessed the efficacy of somatostatin analog (SMS) administration in relieving headaches caused by a pituitary adenoma exceeding 1 cm in size (i.e., macroprolactinoma). A macroprolactinoma typically induces severe headaches through two mechanisms: excessive hormonal secretion and compression of neural adjacent structures. Following a baseline day, alternating doses of fifty micrograms of SMS or placebo were injected in the morning, afternoon, and evening. The patient recorded headache severity using a visual-analog scale ranging from zero (no headache) to one (worst headache ever experienced). Outcome measurements were recorded at three time points: immediately before injection (baseline or pre-injection), one hour post-injection, and three hours post-injection. The authors report: “Compared with the pre-injection headache score, the one-hour post-injection value improved by 55 ±14 percent (mean ±SEM) after SMS but worsened markedly after four injections of placebo.” The original data figure is presented in **Figure S3**. The reconstructed temporal data is plotted in **Figure S4**. The primary outcome measure is the relative difference in headache score compared to the pre-injection score.

The delta headache rating was calculated based on headache rating post-injection related to verum and placebo (**Figure S4**, right panel). The median SMS-induced pain reduction, relative to the placebo, was calculated and plotted per day based on a sequentially updated standard Bayesian regression model on the delta headache rating. The model included priors to constrain parameter estimates to clinically meaningful ranges. The intercept was assigned a Gamma distribution prior (shape: 2, rate: 3), restricted to the range [0, 1], while the slope for the period predictor was assigned a Normal distribution prior (μ: 0, σ: 5), bounded between −1 and 1. The model was fit with 25,000 total iterations, including a 3,000-iteration warmup.

The logarithmic Bayes factors for SMS-induced pain reduction (H1) versus no difference between SMS- and placebo-induced pain reduction (H0) was calculated based on calculated posterior probabilities.

### 2c. Re-evaluating brain stimulation in epilepsy

A third N-of-1 trial assessed the therapeutic efficacy of hippocampal electrical stimulation in four patients diagnosed with refractory mesial temporal lobe epilepsy (Tellez-Zenteno et al., 2006). The longitudinal outcome data was presented for a single case spanning four consecutive years of electrical stimulation intervention. A double-masked design with multiple crossovers was employed, implementing a randomized controlled study protocol comprising three cycles of paired monthly treatment periods. Within each treatment pair, the stimulation device was subjected to randomized activation for one month followed by a month of deactivation. The investigation established monthly seizure frequency as the primary outcome measure. The investigators concluded: “Hippocampal stimulation produced a median reduction in seizures of 15%. All but one patient’s seizures improved; however, the results did not reach significance.” The double-blind, randomized crossover N-of-1 trial was subsequently complemented by an extended open-label observation period for a single patient (**Figure 4**).

**Figure 4.**
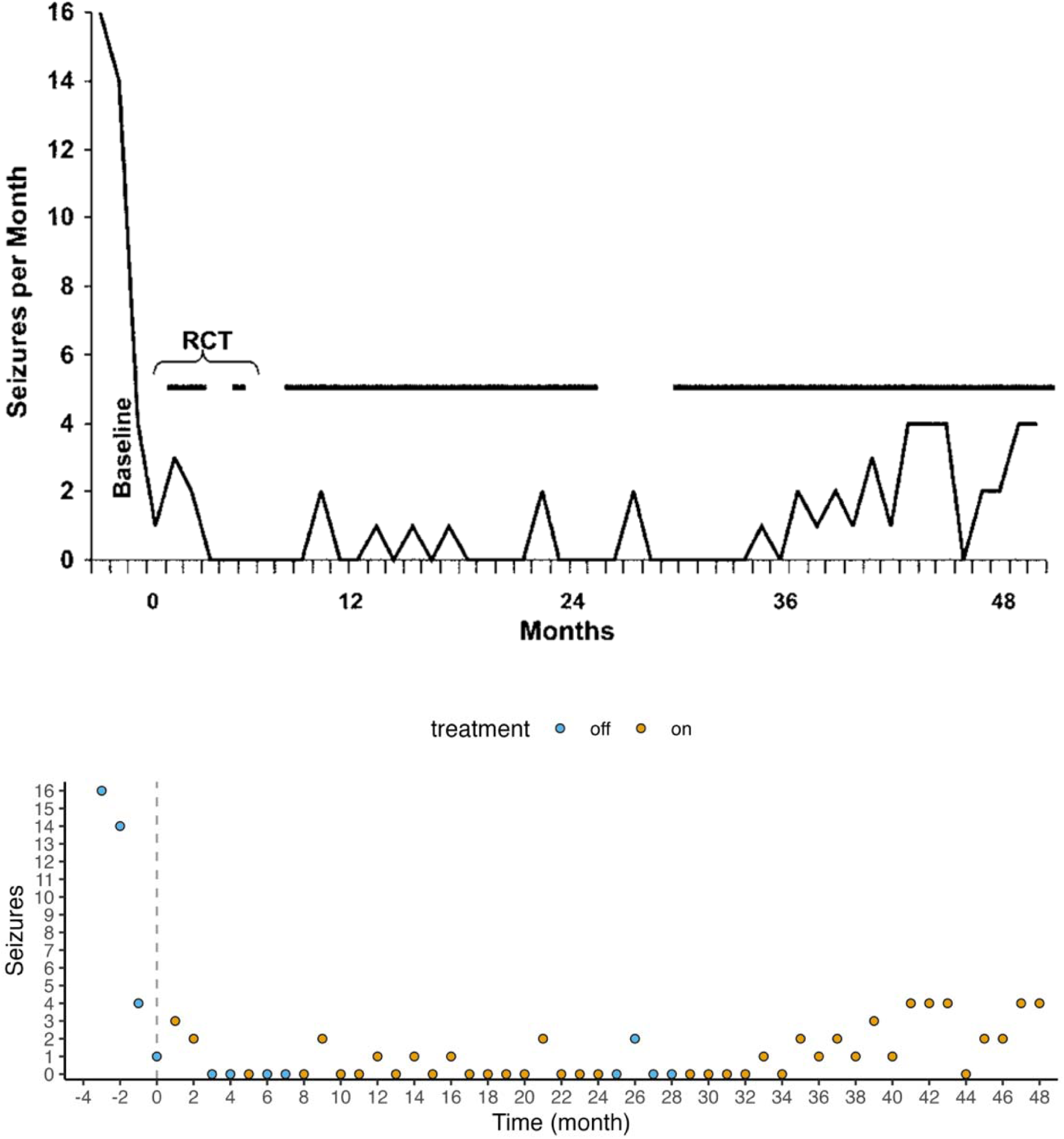
The original monthly seizure representation for four years of consecutive follow-up after an initial double-masked, randomized N-of-1 trial day (RCT) with the electrical stimulator on (black horizontal line) or off (no line). Tellez-Zenteno et al. (2006) provided the data from one of the four included subjects. The reconstructed data is shown below, with (corrected) x-axis labels. Months zero to six are part of the N-of-1 trial, each dot represents a period.

For re-analysis of this N-of-1 trial we quantified the reduction in seizure frequency over time for the single patient with temporal data available (Tellez-Zenteno et al. 2006). Results reported by Tellez-Zenteno et al. 2006 showed a net effect of 33% seizure frequency reduction comparing periods with and without hippocampal stimulation to baseline during the N-of-1 trial (Tellez-Zenteno et al. 2006). We fit the monthly seizures to a Bayesian Poisson regression model, initiated with non-informative default priors, and updated the model after each month, starting at month one.

We estimated median seizure reduction per month with its 95% credibility interval. **Figure 5**). Priors were defined to reflect prior knowledge about parameter values. The intercept was assigned a Normal distribution prior (μ: 0, σ: 100) with bounds between −10 and 10, while the treatment effect was given a Normal distribution prior (µ: 0, a: log(2) / 2): these priors ensure that 95% of the posterior probability lay within a range corresponding to a rate reduction of half to double. The model was fit with 25,000 total iterations, including a 3,000-iteration warmup.

**Figure 5.**
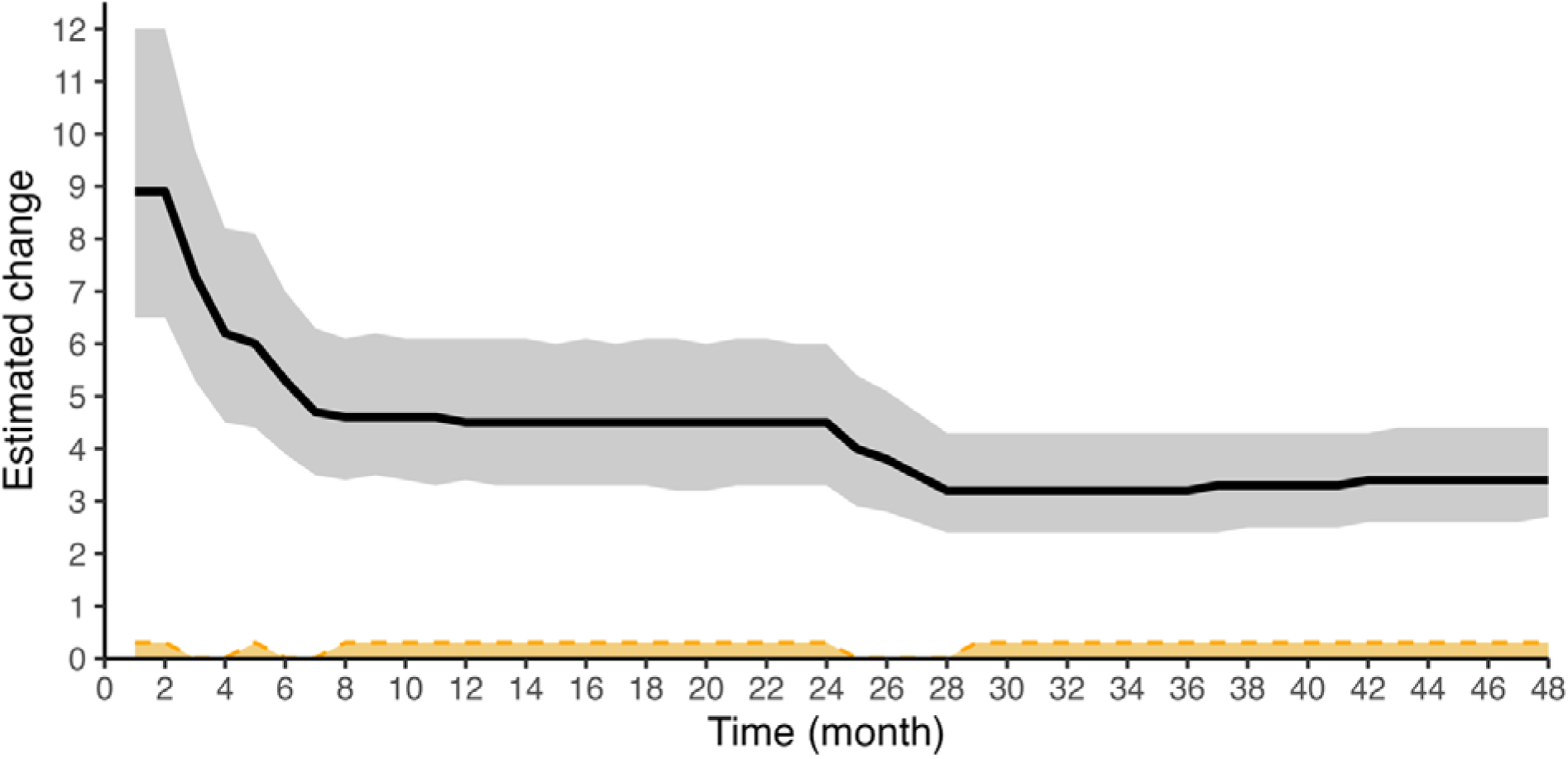
The estimated change in seizure frequency and 95% credibility interval (black line with gray shading) of monthly seizures due to hippocampal stimulation (in orange, stimulation on) over time. The original data by Tellez-Zenteno et al. (2006) is shown in Figure 4. Changes in the estimate and uncertainty occur predominantly when new information is provided during period switching (i.e., on-off transitions). The estimated change (or magnitude of difference) between periods becomes smaller after the last ‘off’ period showing an increase in seizures (24-28 months, Figure 4) reflecting less of a difference in seizure frequency during on-off transitions.

We calculated the logarithmic Bayes factor for a successful seizure reduction due to hippocampal simulation (H1) relative to no stimulation effect (H0) at six, twelve, 24, 28 and 48 months of trial duration.

## Results

### 1. Simulated N-of-1 trial in epilepsy

The simulated data for a representative refractory epilepsy patient (daily seizure rate 5 per day, net effect size 0.5) is shown in **Figure 1**. The first modeling moment is day 85, with at least a single outcome recording for both treatment periods. The H1 model is updated each day until the end of the N-of-1 trial (day 336). **Figure 2** shows the certainty intervals over time for these days based on the posterior probabilities extracted from the H1 model.

After each cycle completion, the H1 model is formally compared to an H0 model. The evidence in favor of H1 over H0 (i.e., logarithmic Bayes factor) increases incrementally from 4.9 (after cycle 1, day 112), 19.9 (after cycle 2, day 168), 33.6 (cycle 3, day 224), and 43.2 (cycle 4, day 280) to ultimately 60.9 after study completion (day 336). After cycle 1 (day 112), there is ‘very strong evidence’ in favor of a difference in treatment effects between verum and placebo periods (H1, alternative hypothesis). However, the Bayes Factor does not provide information on the magnitude of treatment effect – that is: has a meaningful seizure frequency reduction (e.g., 30%) been achieved?

The simulated N-of-1 trial incorporates an MCID analysis of a seizure frequency reduction of 30%. The MCID threshold is graphically represented in **Figure 2** as a horizontal demarcation at –1.5 (denoting a reduction of 1.5 seizures). Sufficient empirical evidence for achieving the MCID is established when the credibility interval exhibits three characteristics: narrowing in range, directional alignment with the MCID, and exclusion of the MCID threshold value. To elaborate, there exists a 95% probability that the true treatment effect estimate, conditional upon the observed data, falls within the boundaries of the 95% credibility interval. When the lower bound of the credibility interval exceeds the MCID threshold in the negative direction, this indicates an increased probability of not achieving the clinically significant reduction. Conversely, once the credibility interval encompasses values equal to or surpassing the MCID, we can reasonably infer that the estimated treatment effect exceeds the predetermined threshold value, given the available empirical evidence.

At day 152 (second cycle), the analysis demonstrates sufficient evidence for an MCID in seizure reduction (median daily seizure reduction: 2.21, 95% credibility interval: 1.51 to 2.92; **Figure 2**). Analysis conducted at this specific day yields an estimated logarithmic Bayes factor would be 15.0, indicating ‘extreme evidence’ supporting the alternative hypothesis (H1). Consequently, utilizing continuous data analysis methodology, the N-of-1 trial could be terminated at day 152 rather than day 168, resulting in a trial duration reduction of 16 days.

A noteworthy observation emerges from this simulated trial: while the Bayes factor at the conclusion of cycle 1 (day 112) suggests “very strong evidence in favor of H1” (4.9), the certainty of achieving meaningful clinical effects remains limited during cycle 1 and portions of cycle 2. During this period, both the median daily seizure reduction and its corresponding certainty interval exhibit considerable breadth and temporal fluctuations as additional data accumulates. An additional 40-day observation period proves necessary to establish temporal stability of effects and to conclude the trial upon achieving meaningful clinical effect. This temporal requirement illustrates a critical methodological distinction: while sufficient evidence supporting the alternative hypothesis (H1) exists at cycle 1’s conclusion, establishing certainty of clinically relevant effects necessitates additional longitudinal data points.

### 2a. Re-evaluating previous N-of-1 trial (Willoughby et al., 2003)

We updated the model fitted to Willoughby’s data after each day, starting on the first day seizures were present (i.e., day 22). The total absence of seizures in the nicotine period (see previous **Figure 3**) and multiple days of seizures during placebo administration results in a probability of direction exceeding 99% achieving convergence to unity within a five-week period (**Figure 6**).

**Figure 6.**
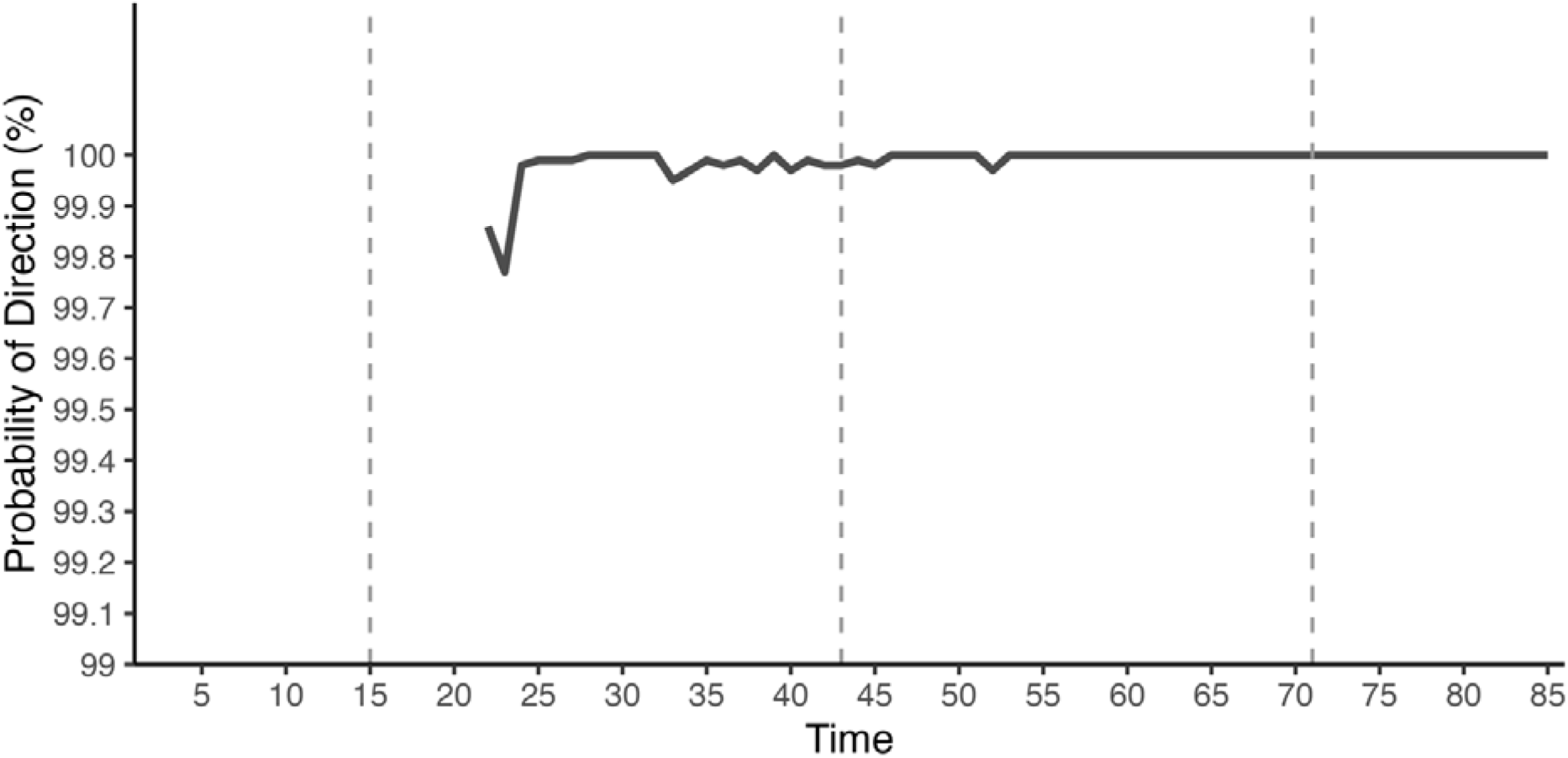
The probability of direction for the existence of a seizure reduction effect using nicotine patches is given over time for the N-of-1 trial daily seizures reported by Willoughby et al., 2003 (Figure 3). The model is initiated on day 22, the first moment the seizures occur, and updated day by day. The certainty of direction is very high and reaches a stable 100% as soon as day 52.

Model initialization occurred on day 22, coincident with the first documented seizure event, followed by systematic daily updates. Directional certainty demonstrates remarkable robustness, achieving and maintaining complete certainty (100%) by day 52 (during cycle 2). The logarithmic Bayes factor, evaluating the relative evidence for H1 versus H0, demonstrates progressive strengthening from 4.3 post-cycle 1 (day 28) to 18.0 following cycle 3 (day 84).

Given that directional probability analysis operates independent of the null hypothesis of treatment inefficacy, the integration of these findings with Bayesian factor analysis – incorporating the relative evidence against the null hypothesis – provides a more comprehensive interpretative framework. Based on this integrated analytical approach, the N-of-1 trial could have been definitively terminated at day 52, representing a potential reduction in trial duration of 32 days (approximately one month) compared to the original protocol.

### 2b. Re-evaluating previous N-of-1 trial (headache)

Analysis of the multi-day consecutive N-of-1 trial conducted by Williams et al., 1986 reveals progressive enhancement in the certainty of SMS-induced pain reduction (**Figure 7**). The initial phase, encompassing the first and second injection days, demonstrates inclusivity of zero within the 95% credibility interval of the SMS-placebo differential in headache score reduction. However, the longitudinal progression through days 3 and 4, incorporating cumulative verum and placebo administration, manifests a substantial verum-induced pain reduction – relative to placebo effect – of 0.45, with uncertainty boundaries delineated at 0.23 and 0.68 upon conclusion of day four. Notably, a significant SMS-induced effect manifests on the penultimate day, albeit with diminished magnitude.

**Figure 7.**
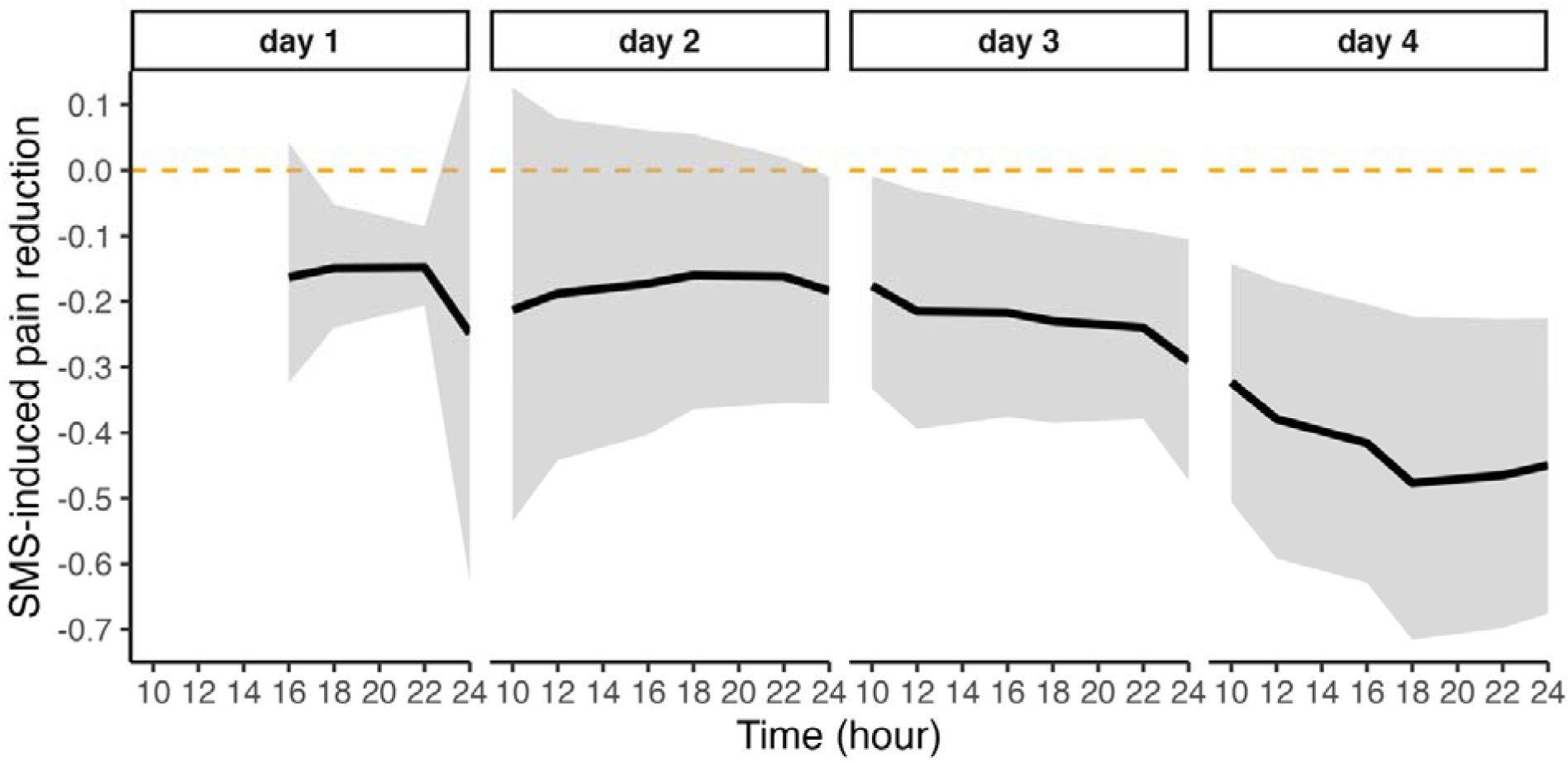
The median SMS-induced pain reduction, relative to the placebo, is plotted over four days based on a sequentially updated standard Bayesian regression model on the delta headache rating (see Williams et al. (1986), **Figure S4**, right). The shaded area depicts the 95% credibility interval.

The logarithmic Bayes factors, evaluating SMS-induced pain reduction (H1) against equivalent placebo- and SMS-induced pain reduction (H0), demonstrate temporal evolution: −0.44 (day one), −0.22 (day two), 2.1 (day three), and 4.3 (trial completion at day four). While the evidence supporting SMS-induced pain reduction achieves ‘strong’ status, this discriminative power necessitates the complete N-of-1 trial dataset. A clinically meaningful effect emerges at day 3; however, the limited data points necessitate additional observations to establish robust evidence favoring verum-induced effect (**Figure 7**).

### 2c. Re-evaluating previous N-of-1 trial (hippocampal electrical stimulation)

The estimated median seizure reduction per month with its 95% credibility interval is plotted in **Figure 5**. During periods of stable stimulation paradigms, the seizure reduction estimate maintains relative constant; however, paradigmatic alterations precipitate diminished uncertainty through enhanced informational input. A significant paradigm shift occurs between months 24 and 28 (hippocampal stimulation discontinuation) (**Figure 5**), whereupon the median change in seizure frequency demonstrates a quantifiable decrease, accompanied by a constriction of the certainty interval encompassing the median. This refined estimate maintains stability throughout the final 20-month observational period (**Figure 5**), persisting despite the occurrence of breakthrough seizures.

Bayesian analysis of successful seizure frequency reduction reveals temporally fluctuating ‘extreme’ evidence for H1, with logarithmic Bayes factors evolving as follows: 1.3 (six months post-trial), 6.8 (one year), 18.7 (two years), 12.2 (28 months), and 6.1 (four years follow-up). The observed decline in logarithmic Bayes factor at 28 months corresponds directly to the paradigmatic shift in hippocampal stimulation cessation. Two extended periods of continuous hippocampal stimulation are documented (6 to 24 months, 28 to 48 months) (**Figure 5**), characterized by distinct trajectories: the initial period exhibits logarithmic Bayes factor elevation, while the subsequent period demonstrates a reduction from 12.2 to 6.1. These changes in the logarithmic Bayes factor reflect the observed variation in mean seizure frequency reduction. This longitudinal investigation exemplifies the dual impact of intervention alternation and the extended observational duration on treatment effect estimation.

## Discussion

We tested the potential benefits of continuous updating in N-of-1 trials in chronic neurological disorders. Based on a simulated N-of-1 trial in epilepsy, and the re-evaluation of data from three previous studies, we demonstrate that accelerated N-of-1 trial decision-making is possible when outcome estimation is continuously analyzed rather than being deferred to post-cycle evaluations.

The sequential design inherent to N-of-1 trials naturally accommodates regular updating of outcome modeling. Accelerating clinical evidence generation requires new information to be processed probabilistically, incorporating relative model evidence estimation and intuitive visualization of posterior probabilities for the most likely model. While the estimation-visualization approach suffices for N-of-1 trials in clinical practice (Kravitz et al. 2013), evaluating the likelihood ratio against a null model prevents presumptive acceptance of an anticipated treatment effect.

The posterior probabilities of the H1-model align effectively with the N-of-1 trial decision-making process, which progresses towards achieving sufficient certainty for therapeutic decisions. This approach proved successful in both the headache reduction case using an experimental somatostatin analog (SMS) and the application of nicotine patches for sleep-related hypermotor epilepsy. In the former case, while the posterior probabilities of the default model indicated an effect after three days, the evidence against the null model remained insufficient for definitive conclusions. The evidence supporting an SMS effect became clear with one additional day of data.

Our comprehensive re-analysis of three N-of-1 trials, supplemented by a simulation study employing continuous data analysis, demonstrates several significant methodological findings; (a) The feasibility of early trial termination when treatment efficacy becomes evident either after or during a treatment cycle, as demonstrated in both the simulated refractory epilepsy case and the Willoughby et al. (2003) study. (b) The substantial value of accumulating additional data points with alternating interventions to reduce uncertainty regarding the estimated treatment effect. This phenomenon is observable both in cases with anticipated treatment effects (Williams et al. 1986) and in studies requiring extended observation periods (Tellez-Zenteno et al. 2006).

The re-analysis demonstrates the distinctive value of diverse analytical approaches within N-of-1 trials using Bayesian methodology. The probability of direction functions as an index of effect existence; however, analogous to the frequentist p-value, it does not quantify the evidence in favor of the null hypothesis. The Bayes factor offers complementary insights through its concurrent evaluation of the alternative, null hypothesis and empirical data, thereby providing a more intuitively interpretable measure of statistical significance.

Notably, our study reveals a methodologically significant finding: the computation of a Bayes factor indicating “extreme evidence” favoring a treatment effect does not necessarily correspond to the achievement of clinically relevant clinical outcomes. This methodological divergence illuminates the inherent limitations of hypothesis testing while underscoring the importance of incorporating clinically meaningful thresholds into analytical frameworks. This is a consideration particularly salient for N-of-1 trials, whose primary objective is the optimization of treatment selection in individual patients. A simulation study using sequential Bayes Factor testing to optimize trial recruitment in clinical studies, exemplifies how to identify and define a clinically meaningful Bayes Factor threshold (Svensson et al. 2021). To our knowledge, no studies using temporal data for continuous outcome estimation with a defined clinically relevant Bayes Factor threshold have been published.

### Importance of continuous data analysis

A sufficient number of measurements and adequate period length are important for enhancing N-of-1 trial precision. However, frequent data sampling and extended trial designs create financial, logistical, and psychosocial barriers to implementing N-of-1 trials. Surveys of physicians and patients report substantial time investment requirements, partially due to trial duration. This is an important challenge in adopting this design in clinical care (Kravitz et al. 2008; Kravitz et al. 2009; Wilmont et al. 2024). A previous survey examining patients’ experiences with N-of-1 trials highlighted that lack of treatment effect or deterioration while on placebo were primary reasons for withdrawing during the first or second cycle (Nikles et al. 2017). In line with this, physicians expressed ethical concerns about subjecting patients to a placebo again when symptom relief has been achieved with treatment (Wilmont et al. 2024; Guyatt et al. 1986). Continuous data analysis allowing accelerated evaluation of the N-of-1 trials can yield statistically substantiated arguments for early trial termination, thereby addressing concerns regarding trial duration, participant dropout rates, and unnecessary exposure to placebo or ineffective verum.

This continuous data analysis methodology is particularly suitable for N-of-1 trials that monitor paroxysmal events or assess symptom severity at regular intervals throughout the study periods, as demonstrated by the included studies. This methodology is less suitable for N-of-1 trials that rely on single measurements obtained solely at the end of each period (e.g., memory, motor skills, language tasks, behavioral problems, quality of life). The case using a visual analogue scale to evaluate headache intensity one hour after SMS injections serves as an illustrative example. In this case, only a single data point is available per period. Although changes in symptom severity can be captured in the analysis, the limited number of data points results in wide certainty intervals, necessitating additional data points before the N-of-1 trial can be terminated.

Outcome measurements (e.g., events, scores) generated through frequent or continuous data collection during a study period are optimal, as they enable the updating of effect estimates with each new data point throughout the treatment period. Each additional data point contributes to refining both the direction and certainty of the estimated treatment effect. This characteristic particularly positions epilepsy and other neurological conditions with recurrent, measurable symptoms as ideal candidates for N-of-1 trials utilizing robust statistical methods to determine treatment effects.

### Methodological considerations in the included studies

The interpretation of results published by Williams et al., warrants particular methodological scrutiny. A potential outcome reporting bias manifests in the progressive increase of the delta (measurement change) over time both placebo and verum conditions, as illustrated in **Figure S4** (right panel). This systematic increase potentially indicates a learning effect, wherein the patient has learned to recognize treatment effects, leading to unblinding or increased nocebo or placebo effect.

Quantitative assessment of the data from Tellez-Zenteno et al. reveals an inverse correlation between seizure frequency differential (Δ) and sequential stimulation modifications, as evidenced by corresponding logarithmic Bayes factors. The study’s methodological constraints, however, introduce significant confounding variables that compromise both trial interpretation and longitudinal observations. During the three-month baseline phase, seizure incidence was n=12, while the N-of-1 trial phase demonstrated n=4 and n=6 seizures during stimulation-on and stimulation-off periods, respectively (**Figure 4**). While the investigators report a 33% reduction in seizure frequency relative to baseline measurements, temporal analysis reveals peak seizure frequency occurred during early baseline measurements, with a declining trend preceding the crossover phase. The investigators did not address potential pre-existing seizure frequency oscillations, which could have warranted an extended baseline period. Computational modeling of baseline seizure frequency’s impact on trial outcomes emphasizes the critical nature of accounting for seizure periodicity and trial duration when establishing baseline parameters (Karoly et al. 2019).

Other factors affecting estimates of treatment effect in the study by Tellez-Zenteno and colleagues are the presence of carryover effects and changes in concomitant medication. A subsequent publication by MacLachan et al. (2010) reported that hippocampal stimulation effects can persist for up to three months, introducing significant complexity into the comparative analysis of ‘on’ and ‘off’ states in N-of-1 trial data.

### Future perspectives

From the patient’s perspective, the rigidity of classical trial design is considered a general limitation. Adaptive trials constitute a methodological framework wherein design parameters undergo systematic modification in response to preliminary outcomes, thereby optimizing resource allocation and minimizing patient burden while maintaining scientific validity. In contrast to traditional clinical trials, N-of-1 trials possess inherent design flexibility that facilitates methodological adaptations while maintaining investigational rigor.

Additional methodological adjustments have been proposed to minimize subject exposure to ineffective verum, specifically through the implementation of interim assessments. These adjustments include: (a) sequential stopping rules, which allows trial to terminate early when clear evidence of treatment effectiveness (or lack thereof) is observed, and (b) response-adaptive randomization, which allocates patients preferentially to treatments with the highest potential benefit (Kravitz et al. 2013; Senanrathne et al. 2020; Shresta & Jain 2021; Myung et al. 2013; van Dongen, Sprengers & Wagenmakers 2023).

Interim assessments have also been employed in N-of-1 trials, both in clinical practice as part of shared decision making and in research contexts where results from multiple N-of-1 trials are aggregated (Guyatt et al. 1986; Stunnenberg et al. 2018). For example, aggregated N-of-1 trials evaluating the effectiveness of mexiletine in patients with non-dystrophic myotonia utilized interim assessments to sequentially terminate N-of-1 trials once sufficient certainty on treatment effect was obtained. This approach allowed some patients to complete only one or two treatment cycles, instead of the planned three or four cycles (Stunnenberg et al. 2018).

Daily data collection through mobile health applications and wearable devices could enhance the precision of treatment estimates and reduce trial duration. The concept of patient-generated health data aligns closely with the objectives of the proposed methodology for continuous data analysis in N-of-1 trials. The participant burden in N-of-1 trials could be minimized through the implementation of mobile health applications and wearable devices for collecting and synthesizing patient-generated health data. Qualitative research on the utilization of patient-generated health data within N-of-1 trials has demonstrated that patients express enthusiasm regarding daily symptom reporting through mobile health applications (Nikles et al. 2005; Whitney et al. 2018). A mobile health application for design and conducting N-of-1 trials, StudyU, has been developed (Konigorski et al. 2022) and is currently equipped to collect patient-generated data. StudyU allows individuals to follow their progress in the mobile health application, however, statistical analysis of results is only showed after study completion (Konigorski et al. 2022).

In conditions characterized by fluctuating symptoms over time, treatment adjustments typically follow a “trial and error” approach, potentially resulting in temporary exacerbations of symptom severity. The differentiation between improvements attributable to the natural course of disease versus those resulting from therapeutic intervention remains challenging to establish. Contemporary literature has demonstrated the presence of cyclical patterns in seizure frequency among epilepsy patients (Leguia et al. 2021; Karoly et al. 2018), which influence the estimated responder rates in traditional clinical trials (Karoly et al. 2019; Goldenholz et al. 2023). The alternating crossover periods inherent in N-of-1 trials, when extended over a prolonged duration, can effectively mitigate bias introduced by natural disease fluctuations. Consequently, the N-of-1 trials design demonstrates particular suitability for conditions exhibiting cyclical symptom variations over time. Neither the model employed in the simulated N-of-1 trial nor the models for the data derived from the published N-of-1 trials accounted for seizure periodicity. Future studies could address cyclical symptom fluctuations through the integration of daily wearable device data and continuous data analysis, thereby enhancing the precision of N-of-1 trial effectiveness estimates.

To our knowledge this represents the first documented application of a simplified model for continuous data analysis in N-of-1 trials comparing treatment and placebo periods. Future implementations can incorporate corrections for covariates, carry-over effects, and autocorrelation when such biases are empirically justified. Methodological adaptations could incorporate different informative priors and expand on the selected stopping rules. Our investigation may provide methodological guidance for others conducting N-of-1 trials. All statistical estimations and visual representations are conducted within the open-source R environment. For those preferring script-free analytical approaches, user-friendly software applications suitable for these analyses are readily available. Specifically, the open-source JASP software facilitates comparable regression models through an intuitive graphical user interface (JASP Team, 2024). The methodology of continuous data analysis in N-of-1 trials expedited decision-making based on trial outcomes, supporting the possibility of trial termination during active treatment periods.

## Data Availability

All data produced are available on GitHub: https://github.com/wmotte/cont_out_est_nof1trials

## Supplementary Figures

**Figure S1.**
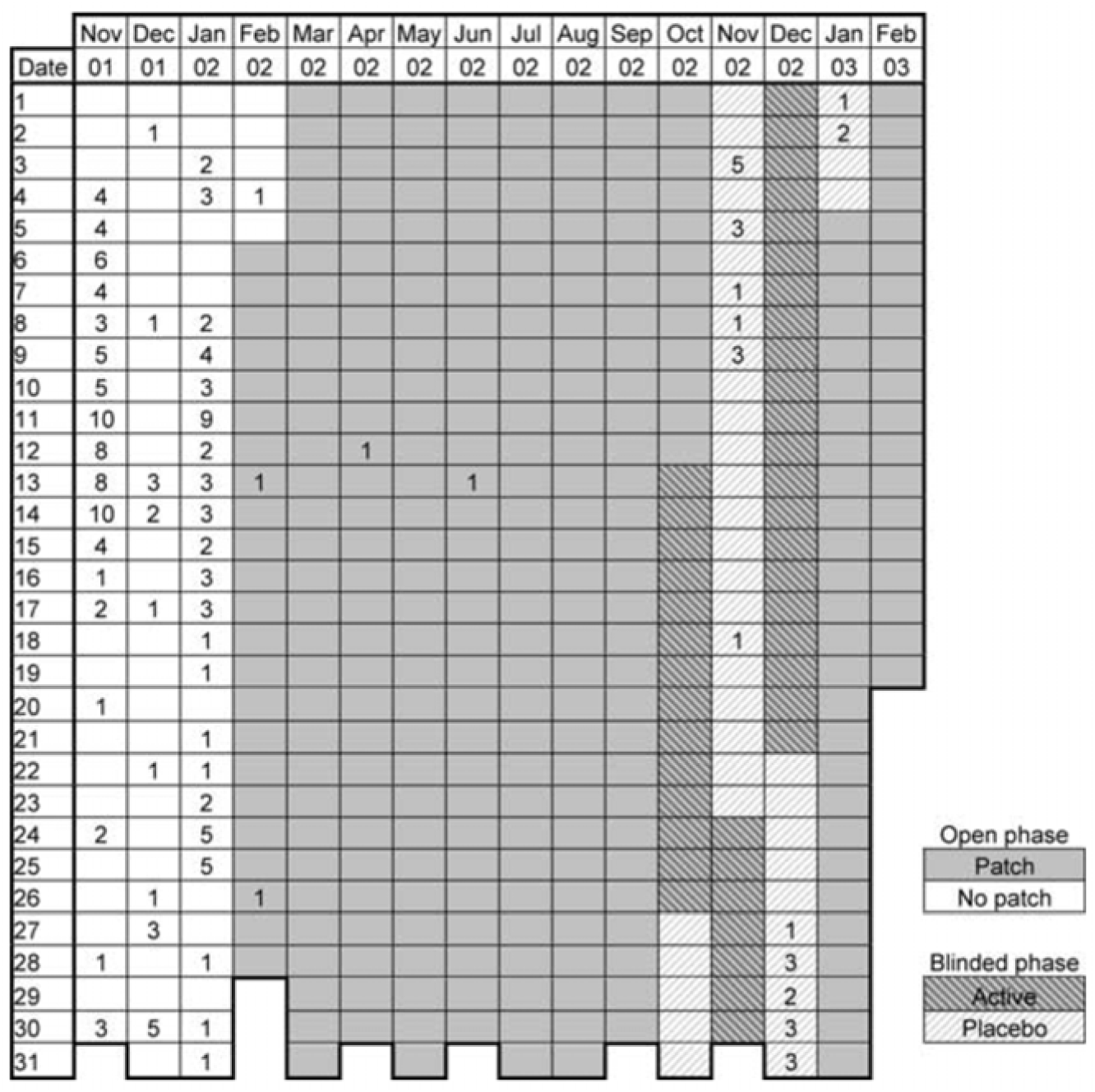
The calendar of seizure occurrences and treatment periods is copied from the Willoughby et al., 2003 paper.

**Figure S2.**
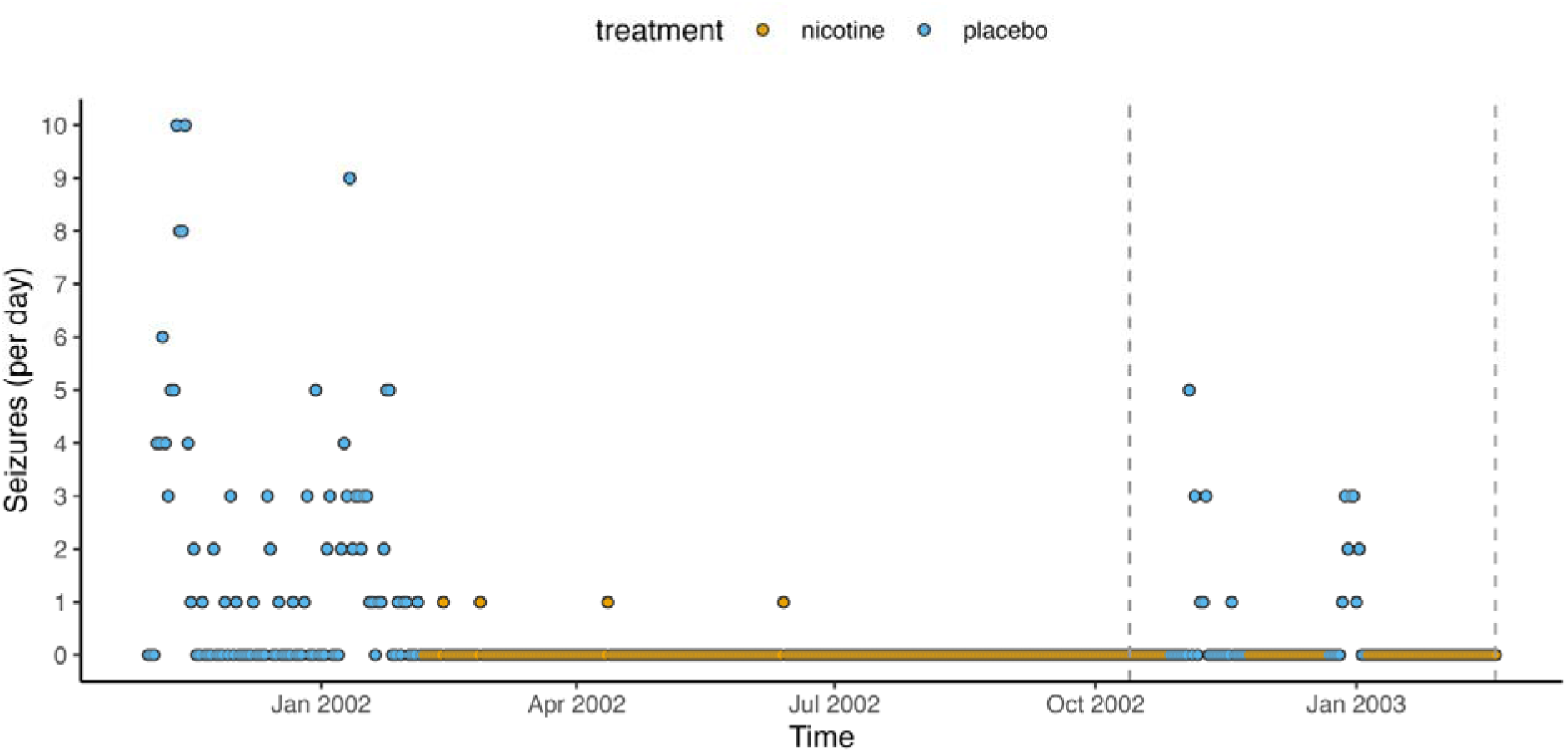
Daily seizure recordings in a single person with refractory epilepsy are reconstructed from Willoughby et al., 2003. A long open-label period is interrupted by a double-masked n-of-1 trial with alternating nicotine and placebo periods of two weeks (indicated with two dashed vertical lines). Most seizures are recorded during the placebo period (highest daily seizure count: ten). Just four seizures are recorded, each on a different day, within the nicotine patch period.

**Figure S3.**
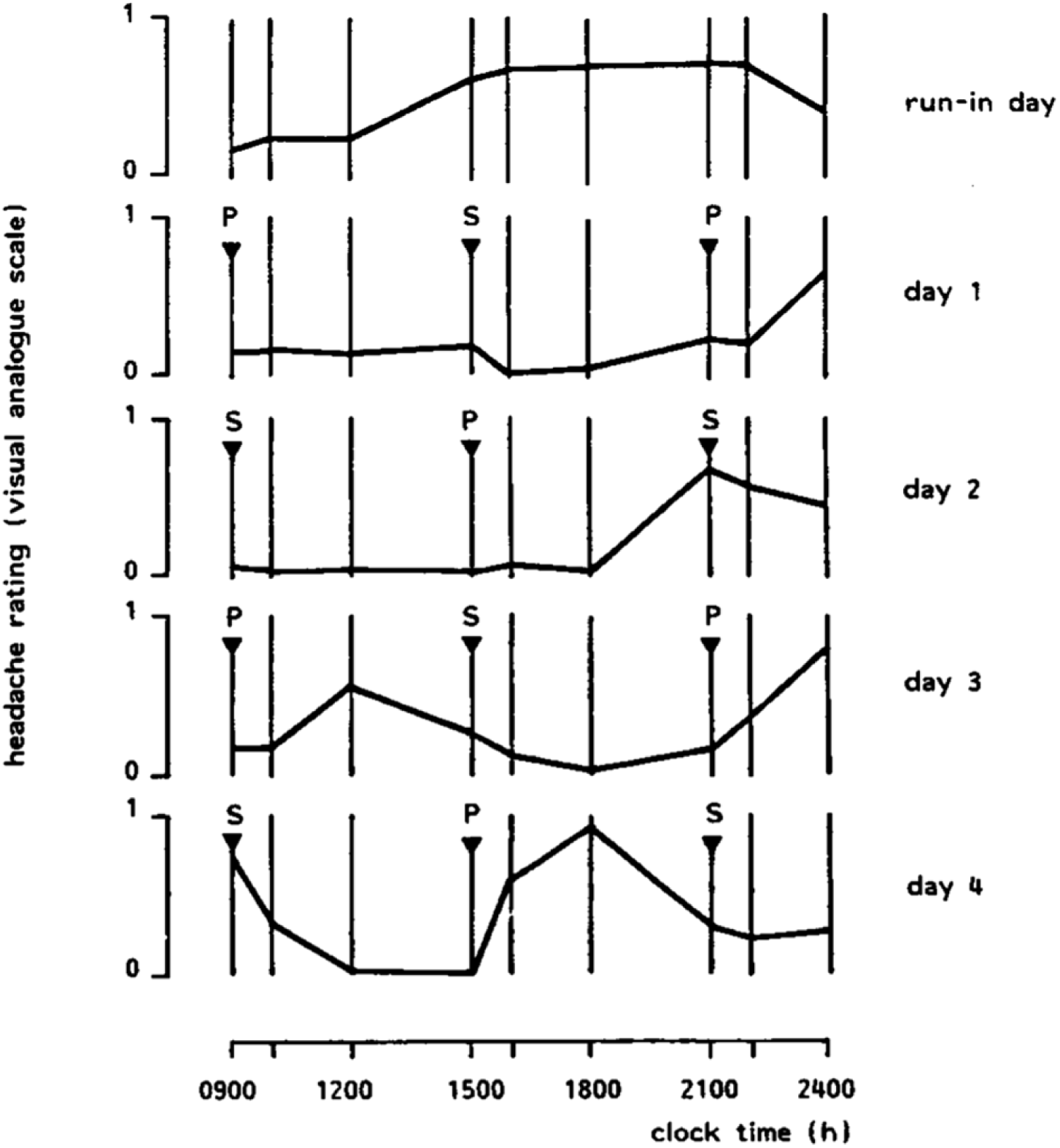
The original visual-analog scoring data for five consecutive N-of-1 trial days was provided by Williams et al., 1986. The first day is without injections. The other four days include an injection at 9 am, 3 pm, and 9 pm with either a placebo (P) or verum SMS 201-995 (S).

**Figure S4.**
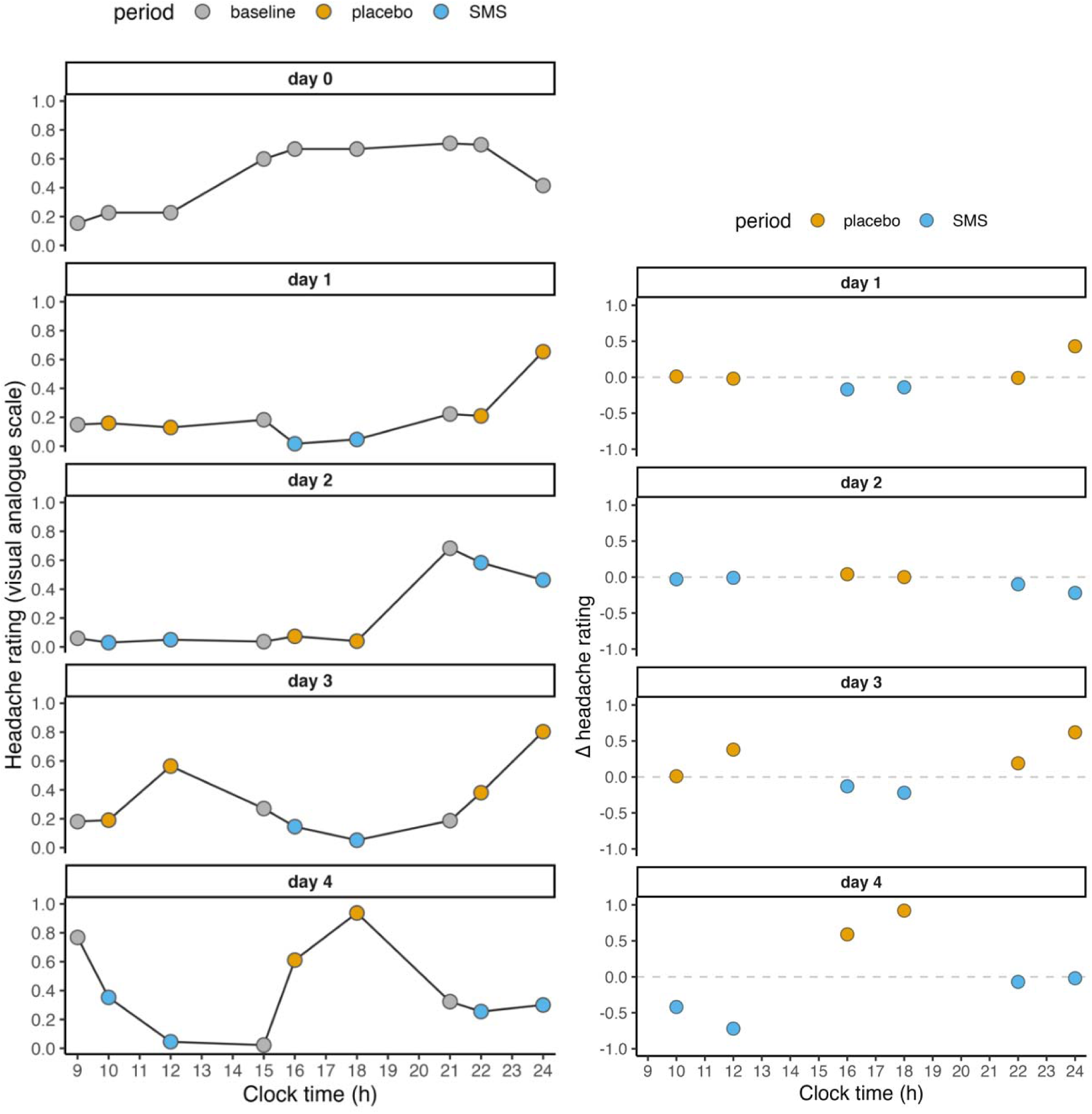
The reconstructed data is based on tracking the original plots for the N-of-1 trial described by Williams et al., 1986 (**left**). The pre-injection times are colored in grey, and the headache scorings at one hour and three hours post-injection are colored in yellow (related to the placebo) and blue (related to the verum; SMS). The outcome of interest is the change relative to the pre-injection. Therefore, the delta headache rating is calculated and plotted alongside the raw rating for the four consecutive days of testing and used as input for the current analysis (**right**).

## Footnotes

1 https://github.com/wmotte/cont_out_est_nof1trials

